# The effect of post-acute rehabilitation setting on 90-day mobility after stroke: A difference-in-difference analysis

**DOI:** 10.1101/2024.01.08.24301026

**Authors:** Margaret A. French, Heather Hayes, Joshua K. Johnson, Daniel L. Young, Ryan T. Roemmich, Preeti Raghavan

## Abstract

**Background:** After discharged from the hospital for acute stroke, individuals typically receive rehabilitation in one of three settings: inpatient rehabilitation facilities (IRFs), skilled nursing facilities (SNFs), or home with community services (i.e., home health or outpatient clinics). The initial setting of post-acute care (i.e., discharge location) is related to mortality and hospital readmission; however, the impact of this setting on the change in functional mobility at 90-days after discharge is still poorly understood. The purpose of this work was to examine the impact of discharge location on the change in functional mobility between hospital discharge and 90-days post-discharge.

**Methods:** In this retrospective cohort study, we used the electronic health record to identify individuals admitted to Johns Hopkins Medicine with an acute stroke and who had measurements of mobility [Activity Measure for Post Acute Care Basic Mobility (AM-PAC BM)] at discharge from the acute hospital and 90-days post-discharge. Individuals were grouped by discharge location (IRF=190 [40%], SNF=103 [22%], Home with community services=182 [(38%]). We compared the change in mobility from time of discharge to 90-days post-discharge in each group using a difference-in-differences analysis and controlling for demographics, clinical characteristics, and social determinants of health.

**Results:** We included 475 individuals (age 64.4 [14.8] years; female: 248 [52.2%]). After adjusting for covariates, individuals who were discharged to an IRF had a significantly greater improvement in AM-PAC BM from time of discharge to 90-days post-discharge compared to individuals discharged to a SNF or home with community services (β=-3.5 (1.4), p=0.01 and β=-8.2 (1.3), p=<0.001, respectively).

**Conclusions:** These findings suggest that the initial post-acute rehabilitation setting impacts the magnitude of functional recovery at 90-days after discharge from the acute hospital. These findings support the need for high-intensity rehabilitation and for policies that facilitate the delivery of high-intensity rehabilitation after stroke.

## Introduction

Nearly 90% of individuals receive rehabilitation during their initial hospitalization for stroke;^8,9^ however, after discharge from the hospital, participation in post-acute rehabilitation is highly variable^10–13^ despite its positive impact on functional outcomes.^1–4^ This variability starts with the initial post-acute setting in which an individual receives rehabilitation (i.e., discharge location from the hospital), with 20-30% of individuals being discharged to an inpatient rehabilitation facility (IRF),^14–17^ 15-25% to a skilled nursing facility (SNF),^14,16,17^ approximately 10% home with community services (i.e., home health—HH—or outpatient clinics—OP),^10,16,17^ and 30-40% home without follow up rehabilitation.^15,16^

Each of these potential discharge locations, or post-acute settings, have distinct requirements that impact the frequency, duration, and intensity of rehabilitation that patients receive. For example, Centers for Medicare and Medicaid Services (CMS) require IRFs to provide at least 3 hours of therapy, 5 days/week; similarly, CMS and other insurers limit the amount of money spent on rehabilitation or the number of rehabilitation visits an individual can receive per year in outpatient clinics.^18^ Due to these requirements and constraints, the frequency, duration, and intensity of care in each of these settings differ, potentially impacting functional outcomes.

Previous work comparing outcomes across discharge locations is incomplete as it focuses on non-functional outcomes or functional outcomes at discharge from one or two post-acute care settings (e.g., IRF only^6,19^ or IRF and SNF^12,20–22^). A majority of prior studies have evaluated non-functional outcomes, such as hospital readmissions or mortality,^6,12,19–21,23^ finding that individuals who are discharged to an IRF have a reduced risk of readmission and mortality compared to individuals discharged to a SNF, although in the short-term IRF care is more expensive.^15^ Surprisingly few studies that have examined the impact of discharge location on functional outcomes, which may impact longer-term costs related to disability. The studies that have examined functional outcomes based on discharge location after stroke have focused on function at time of discharge from IRF and SNF by examining the motor domain of the Functional Independence Measure, finding that individuals discharged to IRFs had greater functional gains during the institutional stay.^24,25^ Due to the limitations of previous work, the impact of discharge location on long-term functional outcomes (i.e., 90 days after discharge from the hospital) remains unclear, yet understanding this is essential for identifying the discharge location that provides optimal patient outcomes at the lowest cost (i.e., high-value of care).

Thus, the purpose of this work is to examine the impact of discharge location after acute stroke on the change in function between hospital discharge and 90-days post-discharge. Here we will focus on functional mobility as it is commonly impaired after stroke, is a frequent target for rehabilitation, and is associated with other health outcomes.^26–29^ We hypothesized that individuals discharged to an IRF would have a larger improvement in functional mobility (as measured by Basic Mobility domain of the Activity Measure for Post-Acute Care [AM-PAC BM]) from the time of hospital discharge to 90-days post discharge compared to individuals discharged to a SNF or home with community services (i.e., HH or OP). This hypothesis is rooted in knowledge of the frequency, duration, and intensity of rehabilitation in each of these settings, where IRF is delivered at the highest frequency and at the lowest frequency in community settings.

## Methods

### Participants

We used data from the electronic health record at Johns Hopkins Medicine in this retrospective cohort study. The Johns Hopkins University’s Institutional Review Board approved the use of these data (IRB00291279). We included individuals over 18 years old who were admitted to either Johns Hopkins Hospital or Johns Hopkins Bayview Medical Center for an acute stroke as defined by ICD10 codes (Supplemental Methods) between July 1, 2016 and December 31, 2022. Individuals were also required to have a measurement of functional mobility via the AM-PAC BM at two time point (see *Functional Measure*). We excluded individuals who were discharged to a location other than an IRF, SNF, or home with community services to ensure accurate categorization of discharge location. Excluded discharge locations included hospice care, long-term care hospitals, cancer centers, and psychiatric centers. Individuals who were discharged home without rehabilitative services were also excluded.

### Exposure

The cohort was stratified into three groups based on the discharge location, which was the exposure of interest. These were IRF, SNF, and home with community services (referred to as Home). The Home group included individuals who were discharged home with either HH or OP rehabilitation. For HH and OP, we specifically examined physical therapy services as mobility is a primary focus of this rehabilitation discipline. This information is documented within a discrete field in the electronic health record. Although we do not have direct information about the dosage of therapy received by individuals at these rehabilitative settings, the settings served as a proxy for rehabilitation dosage early after stroke.

### Functional Measure

We measured mobility with the AM-PAC BM,^30^ which quantifies function on various mobility tasks (e.g., walking, stair climbing). The AM-PAC BM is routinely collected as part of rehabilitation care at Johns Hopkins Hospital and Bayview Medical Center. On inpatient services, the AM-PAC Inpatient Mobility Short Form (commonly referred to as “6-clicks”) is used, while in OP settings the AM-PAC Community Mobility Short Form is used.^31^ The inpatient and community forms are derived from a shared pool of questions,^31^ making T-scores from these separate administrations directly comparable.^32^ Thus, T-scores were used in the analysis.

We used two AM-PAC BM measurements for all individuals. The first measurement was obtained within 48 hours prior to discharge from the hospital (referred to as the discharge measurement) and the second was obtained 60-90 days after discharge (referred to as the 90-day measurement). If an individual had more than one measurement within these separate time windows, we used the measurement closest to discharge or to 90 days, respectively.

### Covariates

In addition to the functional measures and discharge destination, we extracted several covariates from the electronic health record. These covariates can broadly be categorized as demographics, clinical information, and social determinants of health and controlled for disease severity, patient prognosis, and for factors impacting discharge location. Demographic information included age at admission for stroke, sex, race, ethnicity, and the presence (or absence) of select comorbidities. The specific comorbidities were diabetes, hypertension, hyperlipidemia, and a history of depression, which were defined using ICD10 codes (see Supplemental Methods).

Clinical information included hospital length of stay, type of stroke (i.e., ischemic or hemorrhagic) as determined by ICD10 codes (see Supplemental Methods), stay in an intensive care unit (ICU) during the admission (i.e., yes or no), and previous level of function related to mobility (i.e., independent or not independent). We also included information about the amount of physical therapy during the hospital stay, which we defined as the percentage of days after the physical therapy evaluation that the individuals received physical therapy intervention. We accounted for the involvement of other rehabilitation disciplines (i.e., occupational therapy and speech language pathology) with binary indicators. We also included the first AM-PAC BM score from the admission as a covariate to reflect initial severity of mobility deficits. Similarly, we included the first measurement of the Daily Activity domain of the AM-PAC from the admission to reflect initial severity of deficits related to activities of daily living. This metric is routinely collected as part of clinical care and reflects function on various tasks related to activities of daily living (e.g., donning clothes, bathing).^30^

Lastly, we extracted person-and community-level variables related to social determinants of health. The person-level variables were type of insurance at admission (i.e., Medicare, Medicaid, private, or other) and living situation (i.e., living with or without family). The community-level variables were median household income and the Area Deprivation Index (ADI),^33,34^ which is a composite measure of an area’s socioeconomic resources. These metrics were extracted from the American Community Survey and the Neighborhood Atlas,^33^ respectively, and were linked to each patient using census tract information which was derived from the individual’s residential zip code.

### Statistical Analysis

All analyses were conducted in R (v4.0.5).^35^ To examine potential sampling bias, we compared all of the covariates in individuals who were included and excluded in the analysis using Wilcoxon rank sum and χ^2^ tests for continuous and categorical variables, respectively. Similarly, we compared the three groups of individuals included (i.e., IRF, SNF, Home) on covariates and measures of function using one-way ANOVA and χ^2^ tests for continuous and categorical variables, respectively.

To evaluate the impact of discharge location on change in functional mobility, we used a difference-in-differences analysis. Difference-in-differences analyses determine the effect of an exposure—in this case initial discharge location—by examining the differences in the outcome between groups before and after the exposure.^36–38^ The model that was used in this analysis was as follows:

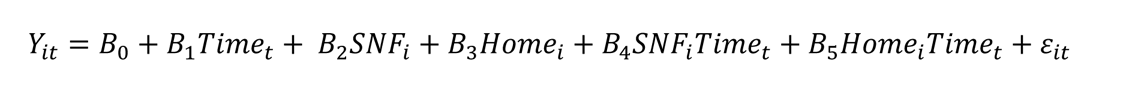

In this equation, *Y*_*it*_ is the AM-PAC BM score at time *t* for individual *i*, *Time_t_* is a variable indicating if the measurement was at discharge (*Time_t_* = 0) or at 90-days post, and *ε*_*it*_ represents covariates. *SNF_i_* and *Home_i_* are variables indicating discharge location of SNF or Home, respectively, resulting in IRF always being coded as 0 and, therefore, serving as the reference group. The coefficient *B*_0_and *B*_1_represent the AM-PAC BM score in the reference group at discharge and the change in AM-PAC BM score from discharge to 90-days in the IRF group, respectively; *B*_2_ and *B*_3_ represent the AM-PAC BM score at discharge in the SNF and Home groups, respectively, compared to the IRF group; *B*_4_and *B*_5_are the difference-in-differences estimators for the SNF and Home groups, respectively, which represents the change in AM-PAC BM score from discharge to 90-days in SNF and Home, respectively, compared to the IRF group. The visualization of the interpretation of these coefficients are shown in Figure S1 of the Supplemental Methods. We used the lmerTest (v 3.3) package^39^ to estimate the difference-in-differences models.

The inclusion of pre and post measurements in difference-in-differences analyses accounts for baseline differences between groups.^38,40^ However, there is an assumption that the outcome of interest changes similarly over time if the exposure did not occur (i.e., the parallel trend assumption).^40–42^ When this assumption is not the case, one must control for that variable. Previous research suggests that there are factors that impact an individual’s change in function over time, making us skeptical of the parallel trend assumption. In these cases, there are three primary ways to account for these factors: 1) inclusion as covariates that interacts with time,^40,42^ 2) propensity score matching,^40,42–44^ and 3) propensity score weighting.^41,45^ We completed the difference-in-differences analysis with and without adjustment. Details about the methods for propensity score matching and weighting are in the Supplemental Methods.

## Results

### Participants

We included 475 individuals in the final analytic cohort (Figure 1). The primary reason for exclusion of individuals was a lack of AM-PAC BM at 90-days. Compared to individuals who were excluded, those who were included tended to be younger, more likely to be Black, had more comorbidities, and had longer stays during their hospital admission. Importantly, there was no significant difference between individuals included and excluded on their functional status at hospital discharge. (Table S1)

**Figure 1.**
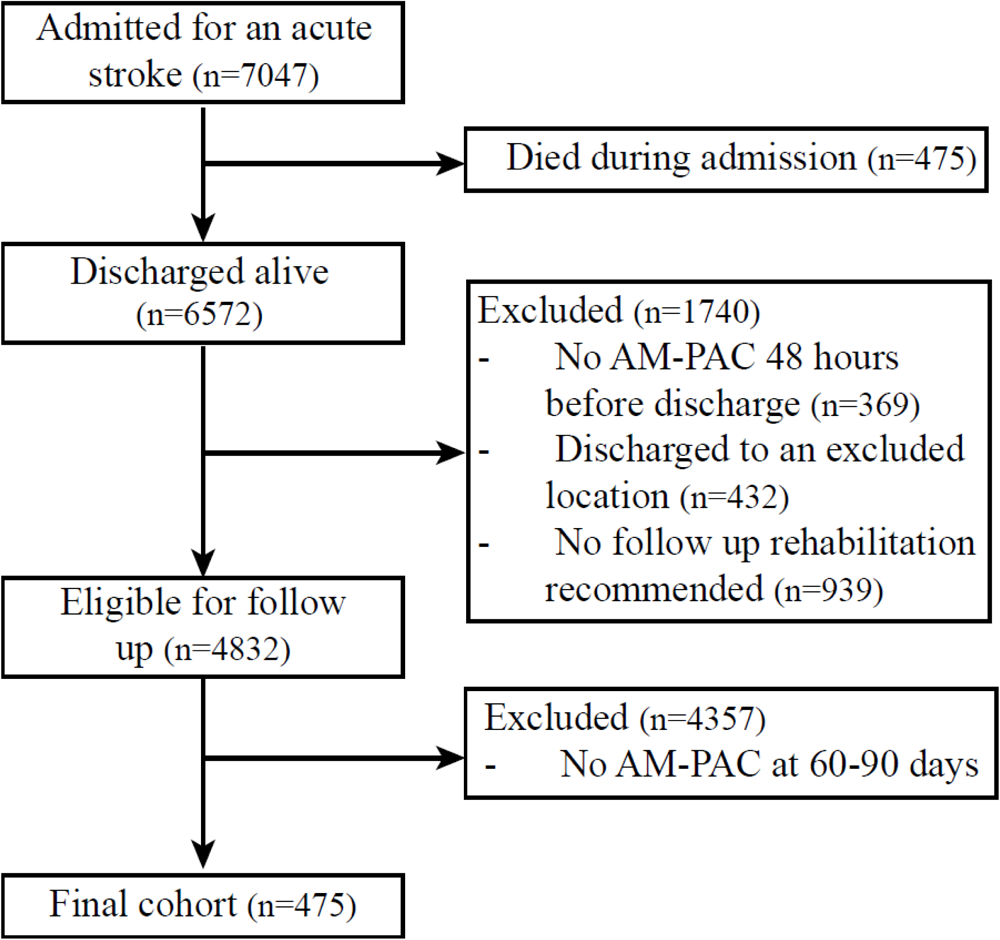
Consort diagram.

Of those included, 182 were discharged home with community services, 103 to a SNF, and 190 to an IRF. Individuals discharged to each of these locations differed on several characteristics, including hospital length of stay, initial functional impairment, insurance, and ADI (Table 1). As described above, a difference-in-differences analysis allows for baseline differences between groups as long as those factors do not impact the change over time,^38,40^ and if this is not the case, then they should be included in in the model as done in the current work.

**Table 1.** Characteristics of individuals by discharge location.

| **Variable** | **IRF\***<br>N=190 | **SNF\***<br>N=103 | **Home with**<br>**community**<br>**services\***<br>N=182 | **p-value<sup>†</sup>** |
| --- | --- | --- | --- | --- |
| **Demographic Information** |  |  |  |  |
| **Age (yrs)** | 63.9 (15.2) | 67.4 (13.7) | 63.4 (15.0) | 0.08 |
| **Sex: Female** | 91 (47.9) | 61 (59.2) | 96 (52.7) | 0.18 |
| **Race** |  |  |  | 0.24 |
| White | 90 (47.4) | 55 (53.4) | 80 (44.0) |  |
| Black | 85 (44.7) | 45 (43.7) | 93 (51.1) |  |
| Other | 15 (7.9) | 3 (2.9) | 9 (4.9) |  |
| **Ethnicity** |  |  |  | 0.78 |
| Not Hispanic | 180 (94.7) | 100 (97.1) | 174 (95.6) |  |
| Hispanic | 7 (3.7) | 3 (2.9) | 6 (3.3) |  |
| Unknown | 3 (1.6) | 0 (0.0) | 2 (1.1) |  |
| **Diabetes: Present** | 79 (41.6) | 51 (49.5) | 80 (44.0) | 0.42 |
| **Depression: Present** | 43 (22.6) | 32 (31.1) | 63 (34.6) | 0.03 |
| **Hyperlipidemia: Present** | 116 (61.1) | 70 (68.0) | 126 (69.2) | 0.21 |
| **Hypertension: Present** | 165 (86.8) | 96 (93.2) | 169 (92.9) | 0.08 |
| **Clinical Information** |  |  |  |  |
| **Type of stroke: Ischemic** | 56 (29.5) | 39 (37.9) | 47 (25.8) | 0.10 |
| **Length of Stay (days)** | 14.3 (17.8) | 20.1 (20.9) | 8.2 (11.4) | <0.001 |
| **Care in the ICU: Present** | 26 (13.7) | 14 (13.6) | 24 (13.2) | 0.99 |
| **Percent days with**<br>**Physical Therapy** | 34.9 (25.0) | 24.0 (20.4) | 42.0 (31.5) | <0.001 |
| **Occupational Therapy**<br>**services utilized: Yes** | 189 (99.5) | 102 (99.0) | 162 (89.0) | <0.001 |
| **Speech Language**<br>**Pathology services**<br>**utilized: Yes** | 152 (80.0) | 70 (68.0) | 89 (48.9) | <0.001 |
| **Initial AM-PAC Basic**<br>**Mobility T Score** | 35.2 (8.4) | 33.5 (8.5) | 44.6 (9.9) | <0.001 |
| **Initial AM-PAC Daily**<br>**Activity T Score** | 32.8 (8.0) | 32.5 (8.3) | 42.2 (9.5) | <0.001 |

**Table 1.** Characteristics of individuals by discharge location
| Variable | IRF*<br>N=190 | SNF*<br>N=103 | Home with<br>community<br>services*<br>N=182 | p-value<sup>†</sup> |
| --- | --- | --- | --- | --- |
| **Previous level of**<br>**function: Independent** | 141 (74.2) | 52 (50.5) | 120 (65.9) | <0.001 |
| **Social Determinants of Health** |  |  |  |  |
| **Living situation:**<br>**Without Family** | 44 (23.2) | 34 (33.0) | 48 (26.4) | 0.19 |
| **Insurance** |  |  |  | 0.09 |
| Medicare | 115 (60.5) | 64 (62.1) | 98 (53.8) |  |
| Medicaid | 21 (11.1) | 20 (19.4) | 33 (18.1) |  |
| Private | 39 (20.5) | 14 (13.6) | 30 (16.5) |  |
| Other | 15 (7.9) | 5 (4.9) | 21 (11.5) |  |
| **Area Deprivation Index**<br>**(national percentile)** | 44.9 (26.0) | 55.0 (24.9) | 52.0 (26.2) | 0.002 |
| **Median household**<br>**income (10,000 US**<br>**dollars)** | 8.2 (4.1) | 6.7 (4.1) | 6.8 (3.8) | 0.001 |
| **Other Information** |  |  |  |  |
| **AM-PAC Mobility T**<br>**Score at discharge** | 39 (7) | 35 (9) | 50 (9) | <0.001 |
| **AM-PAC Mobility T**<br>**Score at 90-days** | 47 (14) | 37(13) | 51 (13) | <0.001 |

Abbreviations: SNF: Skilled Nursing Facility; IRF: Inpatient Rehabilitation Facility; AM-PAC: Activity Measure of Post Acute Care; ICU: Intensive Care Unit
\* Mean (SD); n (%)
<sup>†</sup> One-way ANOVA; Pearson's Chi-squared test

### Difference-in-differences Analysis

The results of the difference-in-differences analysis are presented in Table 2. The results are similar regardless of whether or how covariates were included. Thus, in the text we will discuss only the adjusted model. In the adjusted model, the IRF group was estimated to have an AM-PAC BM score at discharge (β_0_) of 37.4 (3.0), with a change in AM-PAC BM score from discharge to 90-days (β_1_) of 6.3 points (3.3; orange in Figure 2). The SNF group was estimated to have an AM-PAC BM score at discharge that was 2.2 (1.2) points less than the IRF group (β_2_) and a change that was 3.5 (1.4) points less than the change in the IRF group (β_4_). This is interpreted as the SNF group having a lower initial AM-PAC BM score and a smaller change than the IRF group—even after controlling for covariates (Figure 2—purple). The Home group was estimated to have an AM-PAC BM score at discharge that was 7.5 (1.1) points less than the IRF group (β_3_) and a change that was 8.2 (1.3) points less than the IRF group (β_4_). This is interpreted as the Home group having a lower AM-PAC BM score at discharge and a smaller change in mobility function than the IRF group after accounting for covariates (Figure 2— green).

**Table 2.** Summary of four difference-in-differences models where individuals who were discharge to an inpatient rehabilitation facility (IRF) is the reference.

|  |  | Unadjusted (n=475) |  | Adjusted* (n=475) |  | Propensity Matched<sup>†</sup><br>(n=105) |  | Propensity Weighted<sup>‡</sup><br>(n=475) |  |
| --- | --- | --- | --- | --- | --- | --- | --- | --- | --- |
|  | Coefficient | Estimate (SE)<sup>§</sup> | P value | Estimate (SE)<sup>§</sup> | P value | Estimate (SE)<sup>§</sup> | P value | Estimate (SE)<sup>§</sup> | P value |
| **n** |  | 475 |  | 475 |  | 105 |  | 475 |  |
| **Intercept** | $\beta_0$ | 38.7 (0.8) | <0.001 | 37.4 (3.0) | <0.001 | 40.0 (1.9) | <0.001 | 39.2 (0.8) | <0.001 |
| **Time** | $\beta_1$ | 8.0 (0.8) | <0.001 | 6.3 (3.3) | 0.05 | 8.2 (1.6) | <0.001 | 6.5 (0.8) | <0.001 |
| **SNF** | $\beta_2$ | -3.4 (1.3) | 0.01 | -2.2 (1.2) | 0.08 | -3.0 (2.7) | 0.26 | -3.7 (1.3) | 0.005 |
| **Home** | $\beta_3$ | 11.3 (1.1) | <0.001 | 7.5 (1.1) | <0.001 | 9.9 (2.7) | <0.001 | 10.0 (1.2) | <0.001 |
| **Post\***<br>**SNF** | $\beta_4$ | -6.3 (1.4) | <0.001 | -3.5 (1.4) | 0.01 | -3.7 (2.3) | 0.11 | -3.4 (1.1) | 0.002 |
| **Post\***<br>**Home** | $\beta_5$ | -7.3 (1.2) | <0.001 | -8.2 (1.3) | <0.001 | -12.1 (2.3) | <0.001 | -8.2 (1.1) | <0.001 |

\* Age, sex, race, history of diabetes, history of depression, history of hyperlipidemia, history of hypertension, stroke type, length of stay, ICU utilization, previous level of function, initial AM-PAC Basic Mobility and Daily Activity, involvement of occupational therapy and/or speech language pathology, portion of hospital days with physical therapy, living status, insurance type, area deprivation index, and median household income are controlled for in the model.
<sup>†</sup> This model used one-to-one matching on the covariates above, resulting in a reduced sample size.
<sup>‡</sup> In this model, propensity scores were used to weight participants. The above covariates were used to create the propensity score.
<sup>§</sup> Units for these estimates are AM-PAC BM T score points.

**Figure 2.**
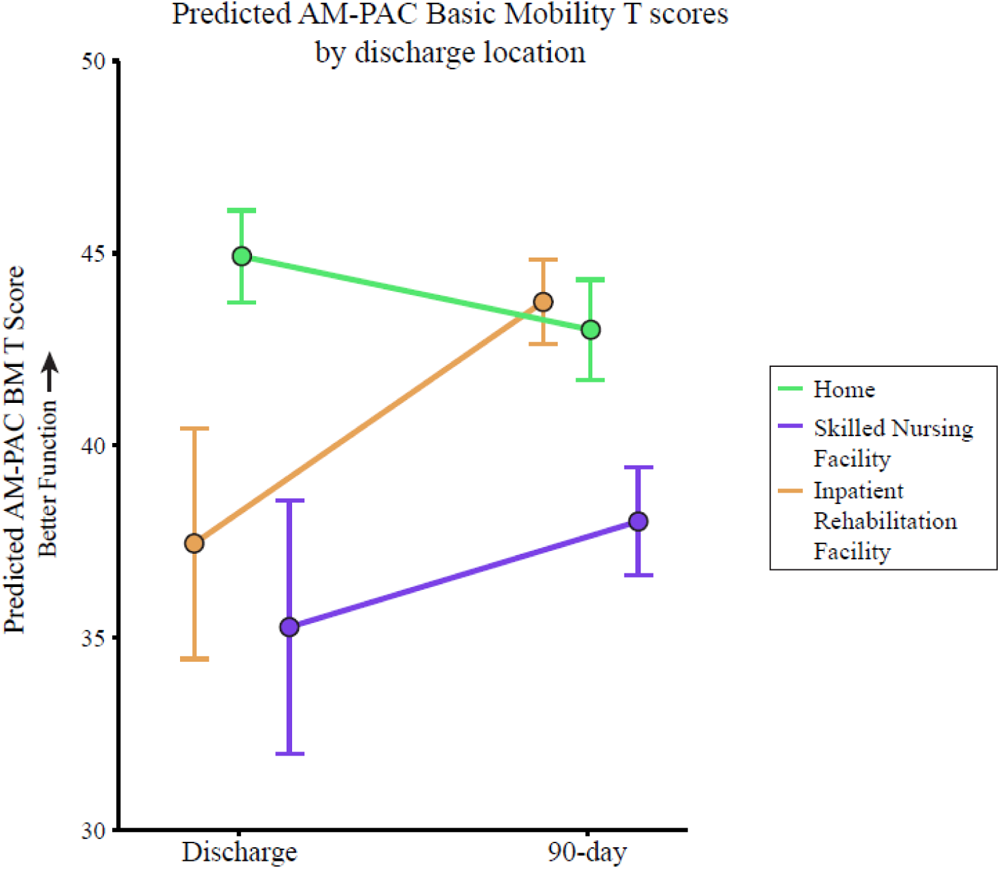
Results of the adjusted difference-in-difference model. Error bars represent standard error.

## Discussion

We examined the impact of discharge location on the change in functional mobility, as measured by the AM-PAC BM, from the time of discharge from the hospital to 90-days after hospital discharge among a cohort of individuals post-stroke. In line with our hypothesis, we found that individuals discharged to an IRF after stroke had a greater improvement in functional mobility than individuals who were discharged to a SNF or home with community services. This was true even after accounting for demographics, clinical information, and social determinants of health using three different statistical approaches. These results add to our current understanding of the impact of discharge location on outcomes by providing insight into the recovery of functional mobility based on discharge location.

Our findings may suggest that we should discharge as many patients as possible to an IRF after stroke due to the higher intensity of rehabilitation. However, the level of intensity is not the only difference in the care provided in each post-acute setting. For example, IRFs involve a more comprehensive team than other post-acute settings, which may not be essential for all patients, depending on their health status at the time of discharge from the hospital. This comprehensive team contributes to higher costs of care in IRFs compared to other settings,^21,24^ creating the need to balance the delivery of high-intensity rehabilitation with the cost of care. Thus, we propose that it may be possible for individuals to obtain similar functional improvement by receiving higher intensity care in lower cost settings. Unfortunately, current reimbursement policies limit the ability to provide high intensity rehabilitation in settings outside of IRF (i.e., HH, OP). Future work is needed to explore the cost-effectiveness of care delivery models that provide higher intensity rehabilitation in non-IRF settings. Critically, this work must be designed to shape policy so that insurers cover high intensity rehabilitation in the most appropriate setting for a given individual.

In addition to policy that supports the delivery of high intensity rehabilitation, policies must be in place to facilitate the measurement of function after discharge from post-acute settings (IRF, SNF, HH, and OP). Current policies (e.g., the Improving Medicare Post-Acute Care Transformation [IMPACT] Act) have standardized quality measures and outcomes during the rehabilitation episode.^46^ However, information on functional status after discharge from post-acute settings is not required. This is beginning to change with the CMS including 90-day functional status and pain as measures of quality of care in patients with a joint replacement under Bundled Payments for Care Improvement (BPCI) and Comprehensive Care for Joint Replacement Model (CJR models).^47^ Similar policies that make systematic measurement of functional status after stroke are necessary to improve outcomes and the value of care.

While our findings suggest that IRFs lead to more improvement in functional outcomes than other post-acute clinical settings (i.e., SNFs and community settings), there is significant variability in the change in functional mobility within each group (Figure 3). The variability supports the need for a shift towards a more individualized understanding of the response to rehabilitation and a more tailored approach to healthcare based on the individual’s characteristics.^48,49^ This approach will allow us to better match individuals with the right care in the right setting to improve the value of rehabilitation. Future work to develop individualized predictive models that can help us understand how a specific individual’s prognosis would change based on the post-acute rehabilitation setting (or the intensity at which care is provided) is needed to reduce this within-setting variability and improve the efficiency of care delivery.

**Figure 3.**
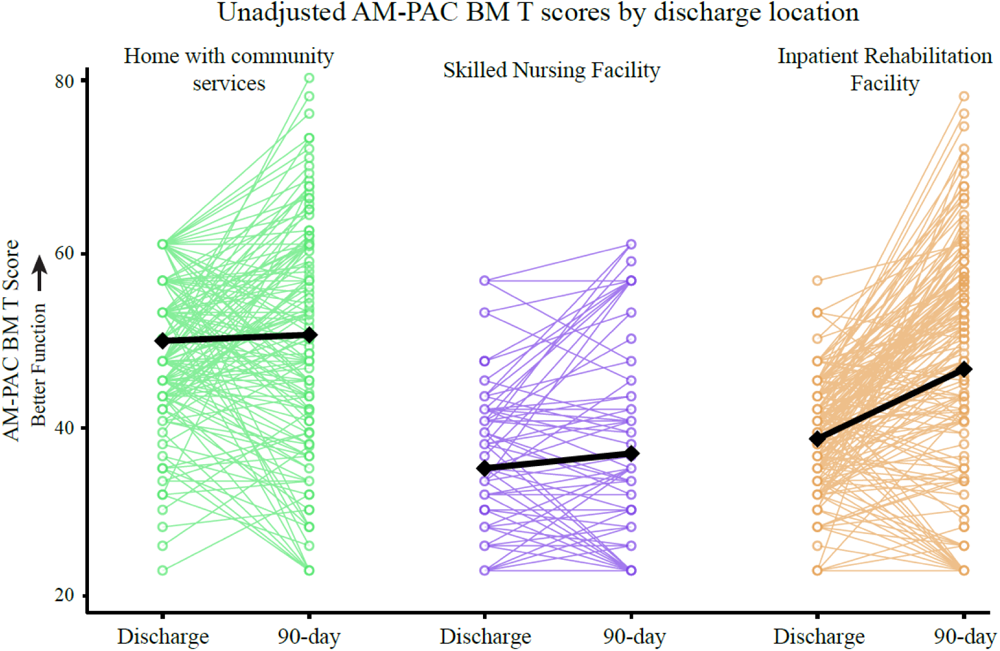
Individual discharge and 90-day AM-PAC BM scores by discharge location. Colored lines indicate individual data while black lines show the group averages.

### Limitations

This work contributes to our understanding of post-acute rehabilitation setting and functional outcomes after stroke; however, it is not without limitations. First, our sample was limited to those who returned to our healthcare system for care because we used electronic health records from a single healthcare system, resulting in the exclusion of a large number of individuals who did not return to our healthcare system. This likely results in the inclusion of more impaired individuals in our analysis. Although this is a common issue in research using electronic health records, it impacts generalizability. Conducting a similar analysis across healthcare systems would overcome this barrier and improve the generalizability of our findings; however, there are many barriers to sharing data across healthcare systems (e.g., patient privacy, data security, standardized functional data collection), highlighting the need for infrastructure to facilitate data sharing across healthcare systems. Additionally, confounding by indication is a concern with this type of analysis. To minimize the impact of this, we controlled for several covariates with three different statistical approaches. Each of these analytical approaches produced similar results, adding to our confidence in the findings; however, as suggested by Simmonds et al (2023),^50^ a clinical trial to examine the impact of discharge location on functional outcomes after stroke would be valuable. Lastly, our measure of functional mobility was the AM-PAC BM, which is a measure that captures one’s perception of difficulty with mobility tasks. While the perception of mobility is an important component of post-stroke recovery, and the AM-PAC BM has good reliability, use of additional measures of functional mobility, such as gait speed and the amount of mobility (e.g., steps per day), would further contribute to our understanding.

### Conclusions

These findings improve our understanding of the impact of discharge location on change in function after stroke. We found that individuals discharged to an IRF had greater changes in functional mobility compared to those discharged to a SNF or home with community services. This lays the foundation for future studies that can inform post-stroke rehabilitation and ensure high value care, where we achieve functional improvement in the optimal clinical setting.

#### Non-standard Abbreviations and Acronyms

IRF: Inpatient rehabilitation facility
SNF: Skilled nursing facility
HH: Home health
OP: Outpatient
AM-PAC BM: Basic Mobility domain of the Activity Measure for Post-Acute Care
ICU: Intensive care unit

## Data Availability

Data is not available in accordance with the institution's policy regarding electronic medical record data; however, code used to in the analysis can be provided upon request.

## Acknowledgments

We would like to acknowledge the Johns Hopkins Precision Rehabilitation Center of Excellence for their assistance in accessing the data for this work and Amit Kumar for his feedback on the manuscript.

## Supplemental Materials

Supplemental methods

Figure S1

Table S1

